# Combined metagenomic- and culture-based approaches to investigate bacterial strain-level associations with medication-controlled mild-moderate atopic dermatitis

**DOI:** 10.1101/2023.05.24.23289041

**Authors:** Nicole M Lane Starr, Numan Al-Rayyan, Jennifer M Smith, Shelby Sandstrom, Mary Hannah Swaney, Rauf Salamzade, Olivia Steidl, Lindsay R Kalan, Anne Marie Singh

## Abstract

**Background:** The skin microbiome is disrupted in atopic dermatitis (AD). Existing research focuses on moderate-severe, unmedicated disease.

**Objective:** Investigate metagenomic- and culture-based bacterial strain-level differences in mild, medicated AD, and the effects these have on human keratinocytes (HK).

**Methods:** Skin swabs from anterior forearms were collected from 20 pediatric participants; 11 participants with AD sampled at lesional and nonlesional sites and 9 age- and sex-matched controls). Participants had primarily mild-moderate AD and maintained medication use. Samples were processed for microbial metagenomic sequencing and bacterial isolation. Isolates identified as *S. aureus* were tested for enterotoxin production. HK cultures were treated with cell free conditioned media from representative *Staphylococcus* species to measure barrier effects.

**Results:** Metagenomic sequencing identified significant differences in microbiome composition between AD and control groups. Differences were seen at the species- and strain-levels for *Staphylococci*, with *S. aureus* only found in AD participants and differences in *S. epidermidis* strains between control and AD swabs. These strains showed differences in toxin gene presence, which was confirmed *in vitro* for *S. aureus* enterotoxins. The strain from the most severe AD participant produced enterotoxin B levels >100-fold higher than the other strains (p<0.001). Strains also displayed differential effects on HK metabolism and barrier function.

**Conclusions:** Strain level differences in toxin genes from *Staphylococcus* strains may explain varying effects on HK, with *S. aureus* and *non-aureus* strains negatively impacting viability and barrier function. These differences are likely important in AD pathogenesis.

**KEY MESSAGES:**

- Staphylococcal strain effects, more so than species effects, impact keratinocyte barrier function and metabolism, suggesting that strain level differences, and not species-level, may be critical in AD pathogenesis.
- The microbiome from mild, medicated atopic dermatitis patients harbor *Staphylococcus* strains with detrimental effects on skin barrier, and may not only be mediated by *S. aureus*.

**CAPSULE SUMMARY:** Patients with mild atopic dermatitis controlled by medication may still harbor strains of *Staphylococcus spp.* that carry toxins that negatively impact skin barrier function.

## INTRODUCTION

Atopic dermatitis (AD) is the most common inflammatory skin disease in children, with recent estimates suggesting as many as 1 in 5 children experience AD, with 80% exhibiting symptoms within the first 6 years of life^1–3^. AD is characterized by recurrent eczematous lesional skin sites. During flares, these erythematous patches are associated with pruritis, exudation, blistering, and lichenification, which can cause loss of sleep, poor mental health, and decreases in school/work performance^1, 5^. It has been proposed that disruptions to the skin barrier during AD flares promote AD pathogenesis^6, 7^. Numerous causes have been proposed for this disruption of the skin barrier, including genetic mutations (for example, in the filaggrin gene)^8^, immune dysregulation (classically characterized by Th2 and IgE predominance)^4, 6, 7^, environmental triggers (such as detergents and epicutaneous protease exposure)^9–11^, and alterations in the skin microbiome (particularly overgrowth of *Staphylococcus aureus*)^12, 13^.

The microbiota plays a critical role in the maturation and function of the skin barrier^14^, including pH balance^15–17^, water retention^18^, keratinocyte differentiation^19, 20^, wound healing^21–24^, innate^25, 26^ and adaptive immune responses^16, 27, 28^, and pathogen competition^29–31^. Commensal skin bacteria, such as *Staphylococcus epidermidis*, produce antimicrobial peptides (AMPs) which are bactericidal and increase keratinocyte production of host AMPs^29, 32, 33^. These bacterial AMPs are active against *S. aureus* and are depleted on AD skin^29^. In contrast, *S. aureus* is important in AD pathogenesis. Multiple studies have shown an increased prevalence of *S. aureus* on the skin of AD patients, particularly on lesional sites^34, 35^. *S. aureus* is known to produce a number of toxins, with staphylococcal enterotoxin B (SEB) in particular associated with more severe AD^36, 37^. However, existing studies, including those associating increased *S. aureus* with lesional skin, have largely focused on moderate to severe participants while holding AD medications, and have focused on the microbiome at the genus and species levels. Whether *S. aureus* plays a similarly important role in less severe or well-controlled disease remains less well understood.

Additionally, variability at the strain-level in AD progression is only beginning to be addressed, with early studies suggesting strains from severe AD subjects more negatively impact the skin in a murine model^35, 38^.

## METHODS

### Overall approach

Skin swabs were collected from 20 participants aged 0.5-14 years for metagenmic analysis of the microbiome. When present, lesional samples were collected in addition to matched nonlesional samples. In contrast to prior studies, AD participants were primarily mild-moderate and continued use of topical therapies to control disease, mimicking real-world clinical scenarios.

Shotgun metagenomic and culture-based approaches were used to evaluate the microbial communities associated with these samples. To determine the effects of strain variation on keratinocyte viability and barrier integrity, keratinocyte cultures were exposed to conditioned media containing secreted metabolites from select *Staphylococcus* isolates derived from participants with a range of clinical presentations.

### Patient recruitment and classification

Recruitment occurred at the University of Wisconsin (UW)—Madison’s Pediatric Allergy-Immunology and Dermatology clinics. Consent for the study and publication of results was obtained from participants’ legal representatives. Approval was granted by the UW School of Medicine and Public Health’s Institutional Review Board. Participants were assigned non-identifying subject IDs for the study. Atopic dermatitis was defined as board certified pediatric allergist or pediatric dermatologist diagnosis with typical signs and symptoms of disease (eczematous dermatitis with typical morphology and distribution) using Hanifan and Reijka criteria. AD severity was determined as previously described using guidelines based criteria^39^. Age and sex matched controls were recruited if they had no current eczematous rash, no prior history of atopic dermatitis, and were not using any topical medication. See Supplemental Methods for criteria for other allergic disease classification.

### Sample collection

Skin swabs were collected from 20 pediatric participants using standard Copan swabs for metagenomic analysis and eSwabs (Copan) to obtain live isolates. Lesional (if present) and nonlesional swabs were collected from all participants. See Supplemental Methods for full details.

### Metagenomic library preparation and analysis

DNA extraction was done as previously described^40^. Samples were sent to University of Minnesota Genomics Center for library preparation and metagenomic sequencing. Resulting FASTQ files were processed using quality filtering, adapter removal, human decontamination, and tandem repeat removal as described previously^40^. Taxonomic classification and abundance estimation was performed using Kraken2 (v2.0.8-beta)^41^ and Bracken (v2.5)^42^. Reads assigned as *Homo sapiens* or that were unclassified at the genus level were filtered out. Decontam (v1.16.0) was then used to identify and remove contaminant reads. See Supplemental Methods for additional details and Supplemental Figure 2 for read counts at each step. StrainGST (v1.3.3)^43^ was used to infer the presence and relative abundance of specific *Staphylococcus* strains, using a set of 230 representative genomes from across the genus^44^. PubMLST was used to determine strain types (ST) for *S. epidermidis*^45^.

### Bacterial isolation and identification

Single isolates were obtained through selective culturing on multiple media types, until single colonies were obtained. Sanger sequencing on the full-length 16S rRNA gene was used for taxonomic classification. See Supplemental Methods for full detail.

### Whole genome sequencing

Bacteria identified as *Staphylococcus* were sent for whole genome sequencing at SeqCenter (Pittsburgh, PA) using the Illumina DNA Prep kit and IDT 10 bp UDI indices, with sequencing performed on an Illumina NextSeq 2000. Demultiplexing, quality control, and adapter trimming was done with bcl-convert (v3.9.3). These genomes were then used to generate a phylogenetic tree with autoMLST in the *de novo* mode and concatenated alignment functions^46^. The resulting tree was visualized using iTOL^47^. Gene-calling and standard-annotation was performed using PROKKA (v1.13)^48^. Toxin genes were identified through DIAMOND BLASTp alignment to the full Virulence Finder Database^49^ download in Jan 2023, retaining the best hit per query based on bitscore with additional filters to retain only alignments exhibiting a maximum E-value of 1e-5, a minimum percent identity of 50%, and minimum query and subject coverages of 70%.

### Generation of staphylococcal cell free supernatants

*Staphylococcus* isolates were grown from stock on trypticase soy agar overnight at 37 °C. Single colonies were then grown in trypticase soy broth overnight at 37 °C with shaking. The following morning, liquid cultures were used to inoculate 50 mL supplemented keratinocyte growth media (see Supplemental Methods for recipe) to a starting OD_600_ of 0.1, followed by incubation at 37 °C with shaking. OD_600_ was read every 2 hr until an OD_600_ of 0.7 was reached (range 0.693- 1.156, average 0.774; typically 3-7 hrs). The cultures were centrifuged at 3220 x g for 10 min. The supernatants were collected and passed through a Watman cellulose Grade 1 filter paper (GE Healthcare Life Sciences), then through a 0.2 µm pore filter. Supernatants were stored at -20 °C until use. Cultures were serially diluted for CFU plating immediately following inoculation and immediately preceding collection to confirm similar CFUs across strains (average of 2.64 *10^9^ CFU/mL).

### *In vitro* enterotoxin assay

*Staphylococcus* isolates from stock were grown on BHI agar overnight at 37 °C. Single colonies were grown in 3 mL BHI liquid culture overnight at 37 °C with shaking. The next day, the cultures were centrifuged at 3500 x g for 5 min. The supernatants were collected and passed through a pore filter (0.2 µm). Enterotoxin levels were then measured using the BioPharm Ridascreen kit according to manufacturer’s instructions.

### MTT assay

Effects of staphylococcal cell free supernatants on neonatal human keratinocyte (HK; American Type Culture Collection (ATCC; Manassas, Virginia)) cell viability were assessed using the 3- (4,5-dimethylthiazol-2-yl)-2,5-diphenyltetrazolium bromide (MTT) colorimetric assay (Roche Diagnostics, Mannheim, Germany) per manufacturer instructions. See Supplemental Methods for full description.

### TEER assay

HKs were also used for the trans-epithelial electrical resistance (TEER) assay. The cells were cultured in HK media on a semipermeable filter insert in a 5% CO_2_ atmosphere at 37 °C for 24 hrs until they reached confluence. They were then switched to a differentiation medium (DM; DMEM medium supplemented with 1.8 mM calcium ion and 4 mM glutamine (Thermo Fisher Scientific)) alone (Control) or DM containing one of the *Staphylococcus* supernatants diluted to 1*10^6^ CFU/mL. The integrity of the HK monolayer was verified by measuring the TEER assay after 2 days of treatment. Three independent replicates were used to calculate the results. The electrical resistance, measured in ohms, of an empty insert was subtracted from that of the insert with cells to yield the resistance of the cells. The electrical resistance was then multiplied by the area of the insert to determine the TEER. Results are expressed as the percentage change in TEER, which reflects the change in resistance after 2 days of treatment normalized to the TEER value of the control (DM alone).

### Statistical analysis

R (version 4.2.0) was used to do all analysis and figure generation. A Kruskal-Wallis test was used to compare select taxonomic levels and Shannon Diversity Index. PERMANOVA testing was used with Bray-Curtis dissimilarity estimates. These calculations were done on relative abundance set to 100% for microbes, bacteria, or *Staphylococcus*, where appropriate. A t-test was used to compare enterotoxin levels. Following a significant ANOVA, pairwise t-tests were used to compare the MTT and TEER effects of the *Staphylococcus* isolates to media controls, with a Bonferroni correction used to account for multiple testing.

## RESULTS

### Clinical characteristics and sample collection

Twenty pediatric participants, aged 5 months to 14 years, were recruited from the University of Wisconsin Pediatric Allergy-Immunology and Dermatology Clinics (Table 1). One control participant was omitted due to having a parent-reported history of atopic dermatitis (AD) but no physician diagnosis and no symptoms of disease. The remaining participants consisted of 8 healthy controls, and 11 with diagnosed AD, primarily mild-moderate in severity. Participants with AD were not asked to halt usage of topical treatments (Supplemental Table 1). Skin swabs collected from participants were classified as control, lesional, or nonlesional and were processed for DNA extraction and metagenomic analysis and untargeted bacterial isolation.

**TABLE 1.** Demographic data for the participants.

| **PARTICIPANTS** | **N (% of total)** |
| --- | --- |
| Total | 19 (100%) |
| Female | 8 (42.11%) |
| **AGE** | **Years** |
| Range | 0.46 - 14.02 |
| Median (IQR) | 3.07 (1.74 – 6.82) |
| **RACE** |  |
| Asian | 2 (10.53%) |
| Black or African American | 2 (10.53%) |
| Other | 1 (5.26%) |
| White | 14 (73.68%) |
| **AD SEVERITY** |  |
| None | 8 (42.11%) |
| Mild | 7 (36.84%) |
| Moderate | 2 (10.53%) |
| Severe | 2 (10.53%) |

### Skin metagenomes from well-controlled AD participants resemble controls

Metagenomic analysis of skin swabs was completed to provide a comprehensive view of the microbiome across participants. After filtering and quality control, the majority of reads (>97%) were identified as Bacteria (Fig 1A). The remaining 3% of reads mapped to either Eukaryota or Viral databases. The dominant bacterial genera were consistent with previous reports of pediatric skin—*Streptococcus*, *Cutibacterium*, *Micrococcus*, *Staphylococcus* (Figure 1B). Of note, the Eukaryota were dominated by *Malassezia restricta*, but there were no significant differences across genera (Supplemental Fig 3A). Interestingly, viral relative abundance was elevated in AD (p_adj_=0.010). This was likely driven by the presence of *Escherichia virus T4*, which was only identified in AD participants (p=0.004) (Supplemental Fig 3B). Molluscum contagiosum virus, which is associated with skin infection, was also present on the skin of several participants, but was not significantly different among the groups (Supplemental Fig 3B).

**FIGURE 1.**
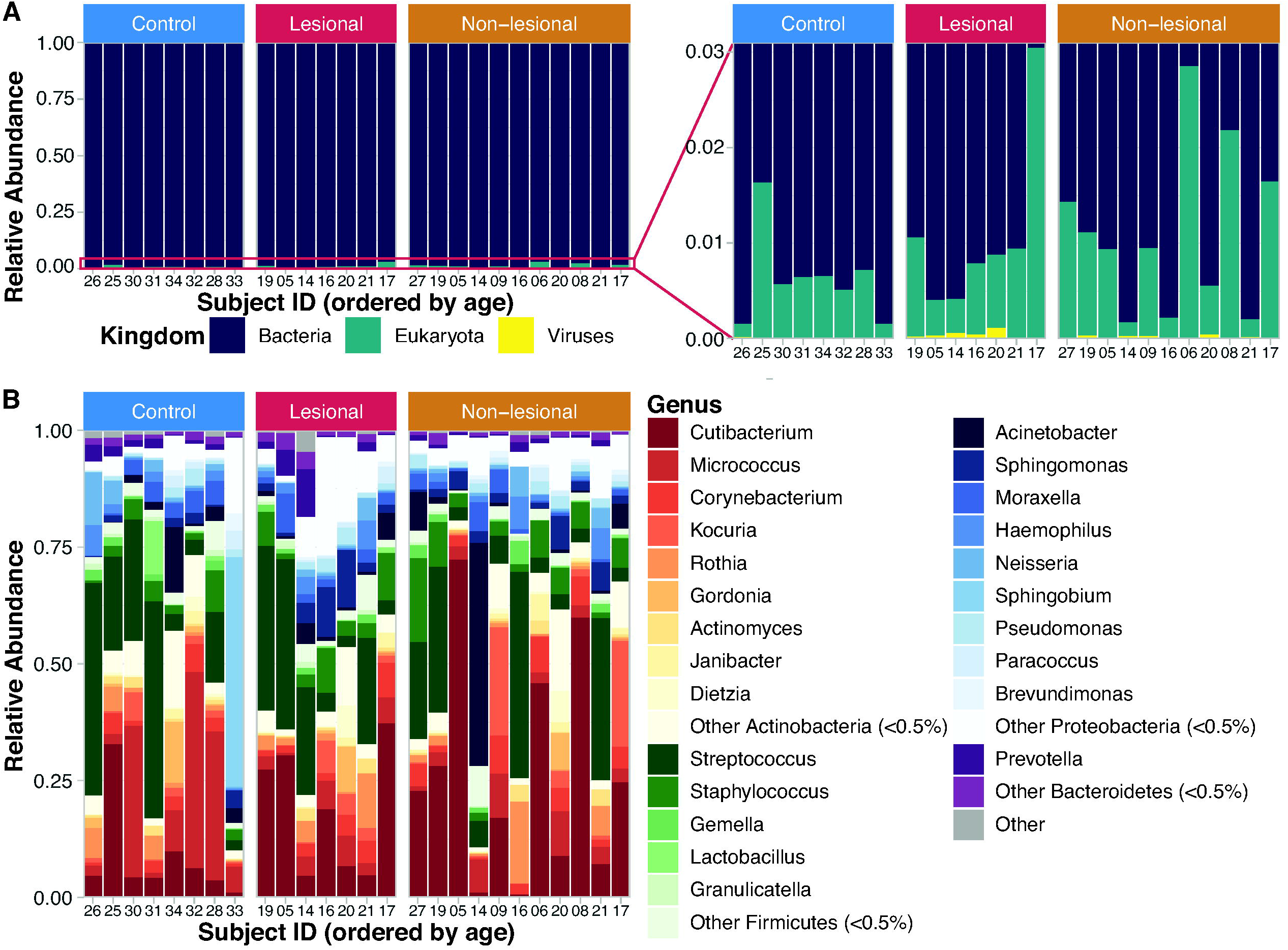
Metagenomic analysis of skin swabs from AD and control groups. Bar graphs of the relative abundances of kingdom-level data (A), with an inset to show differences in Eukaryota and Viruses. Relative abundance plot of bacterial genera (B), with hue corresponding to phylum, genera are ordered by phylum abundance. For both, each swab was normalized to 100% relative abundance of all included taxa along the y-axis and bars are grouped by AD status then ordered by age along the x-axis.

Alpha (within-sample) diversity of the bacteria, as measured by the Shannon Diversity Index, did not significantly differ between the 3 groups (Figure 2A), nor when considering combined AD (pooled lesional and nonlesional samples) vs control samples. However, a trend emerged showing lower diversity but greater evenness in the lesional group compared to control (Supplemental Fig 4). A similar trend was noted with the combined AD group for evenness (p=0.07). Similarly, beta (between-sample) diversity, measured by Bray-Curtis dissimilarity, showed weak grouping of controls vs combined AD (p=0.056).

**FIGURE 2.**
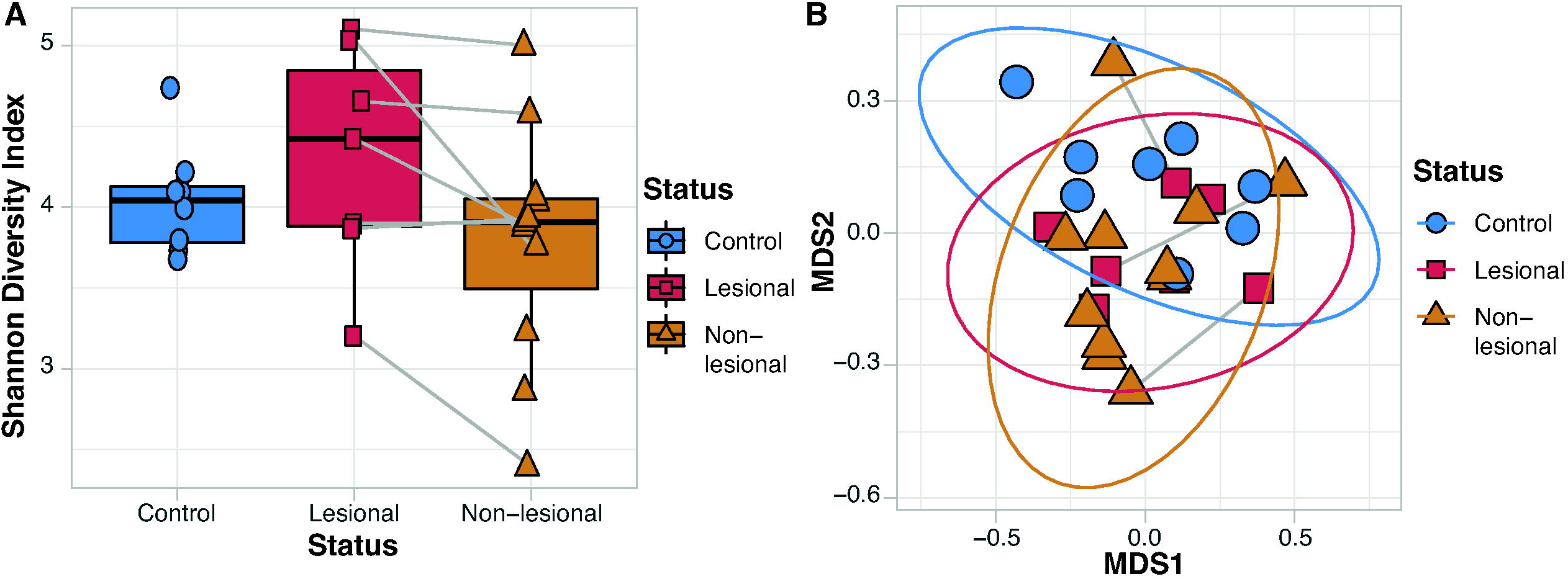
Diversity metrics did not differ significantly by AD status. The Shannon Diversity Index (A) and Bray Curtis Dissimilarity (B) were calculated for all metagenomes and then grouped by AD Status.

### Staphylococcal strain variation by AD status

*Staphylococcus* species are amongst the most dominant taxa on human skin, and blooms of *S. aureus* have been associated with AD flares. Focusing on the *Staphylococcus* species within the metagenomes showed large intra-individual variability (Fig 3A). Although the proportion of *Staphylococcus* species present in the three groups did not significantly differ, the total number of different *Staphylococcus* species present in AD participants trended higher (Fig 3B; p=0.15). Notably, we did find a decrease in Shannon Diversity when comparing combined AD vs control (p=0.048; Fig 3C).

**FIGURE 3.**
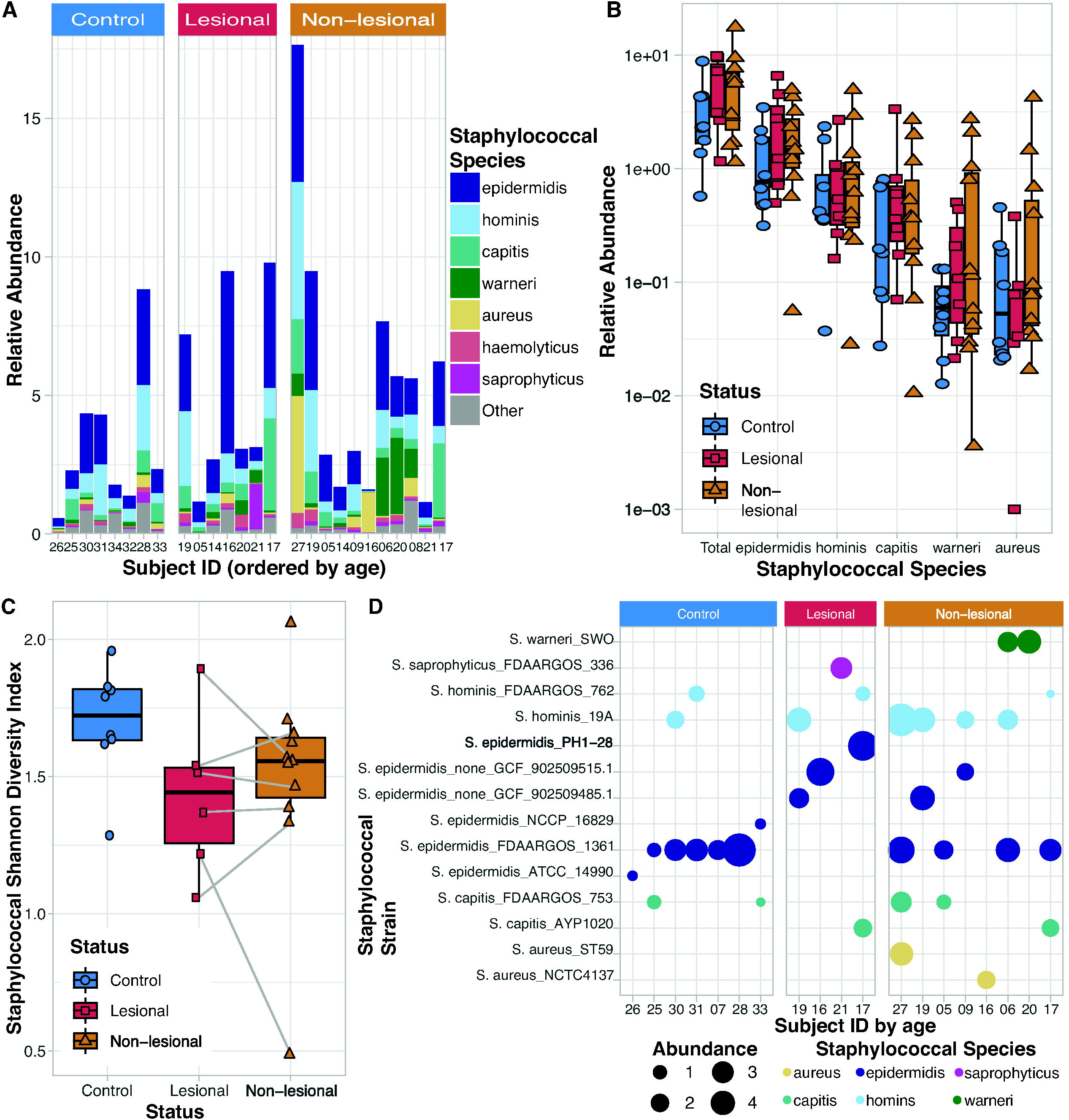
*Staphylococcus* species and strain level variation by AD status. (A) Bar chart showing relative abundances of *Staphylococcus* species detected in each metagenome, grouped by AD status, ordered by age across the x-axis. (B) Bar chart showing the relative abundances of the top 5 *Staphylococcus* species (as called by Kraken/Braken), grouped by AD status. (C) Shannon diversity index of all *Staphylococcus* species (as called by Kraken/Braken), grouped by AD Status. (D) *Staphylococcus* strains (as called by StrainGST), grouped by AD status, ordered by age.

To determine if specific strains of Staphylococci vary by AD status, we classified the *Staphylococcus* strains in metagenomes using StrainGST^43^. This analysis showed a striking lack of *S. epidermidis FDAARGOS_1361* (strain type (ST) 153) in lesional samples, but was present in 5/7 control samples and 4/8 of nonlesional samples (Fig 3D). Conversely, 3 other strains of *S. epidermidis* were present only in AD swabs (ST 5, 89, 387). These data suggest that the association of *S. epidermidis* with disease activity may be strain dependent. Using StrainGST, we observed *S. aureus* strains only in AD samples, specifically in the nonlesional swabs. Strains of *S. warneri* and *S. saprophyticus*, skin commensals that may act opportunistically^50, 51^, were also only observed in AD samples (Fig 3D).

### Isolation and identification of live bacterial isolates

To determine the role different strains may be playing in AD pathogenesis, we performed untargeted bacterial isolation. Using four different media to capture greater microbial diversity, 601 isolates from 26 swabs were obtained. Full length 16S rRNA gene sequencing was used to taxonomically classify each isolate, resulting in the classification of 22 genera—13 among the 305 isolates from control swabs, 10 from the 84 lesional isolates, and 14 from the 210 nonlesional isolates (Supplemental Table 2). Of those, 93% were genera represented in the metagenomes. Actinobacteria were the most abundantly represented phylum, followed by Firmicutes and then Proteobacteria in both the metagenomic- and culture-based approaches. Of note, control swabs had >50% more isolates collected than from nonlesional swabs, which, in turn, had twice the number of isolates as lesional swabs. *Micrococcus* was the most abundantly cultured genera from control samples, while *Staphylococcus spp*. (specifically *aureus*, *capitis*, *epidermidis*, *hominis*, *saprophyticus*, *succinus*, and *warneri*) were the most abundantly isolated in lesional and nonlesional swabs (Supplemental Table 2). Combined, these two genera accounted for >40% of all isolates. Strikingly, the proportion of isolates that are *Staphylococcus spp*. rose greatly in the AD samples, particularly those from nonlesional sites.

### Toxins are elevated in AD-associated Staphylococci

To better understand the role of Staphylococci strain-level differences on AD, we utilized the above-described isolate library. Staphylococci strains representing species isolated across participants were subject to whole genome sequencing. From this, a phylogenetic tree was generated showing relatedness of the selected strains (Figure 4A). Of note, all *S. aureus* isolates were collected from moderate and severe AD participants. Conversely, all *S. epidermidis* and *S. hominis* isolates were from mild AD or control participants. The *S. capitis* strains were split into 2 clusters, one comprised of moderate-severe AD participants and the other associated with a single mild participant. Genes encoding toxin production in each genome were predicted using the Virulence Factor Database. We noted three patterns for genes encoding toxins—those universally present (not shown), those present in a specific species, and those present in only select isolates. All isolates, even coagulase negative Staphylococci (CoNS), contained at least five toxin-associated genes. As expected, *S. aureus* strains contained increased enterotoxin, hemolysin, and leukocidin genes (Fig 4B). Only 1/3 *S. aureus* strains isolated encoded for *seb*. This *S. aureus* strain, designated LK1493, was isolated from a participant recorded as having severe disease. To confirm the genomic predictions, the production of enterotoxins in each *S. aureus* isolate was tested *in vitro*, confirming high levels of SEB production by LK1493 (p<0.001) (Fig 4C).

**FIGURE 4.**
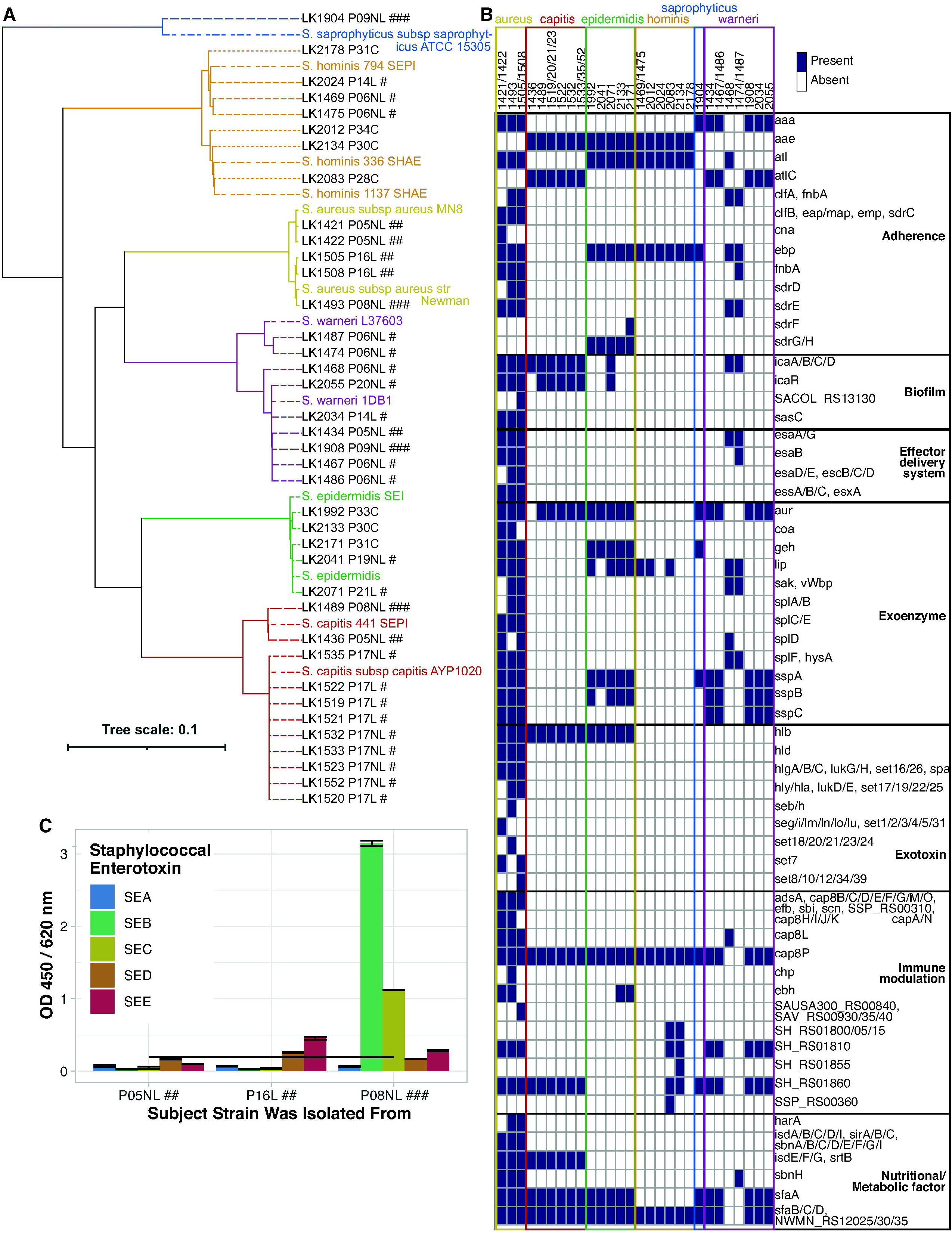
Toxins are elevated in AD-associated Staphylococci. (A) Phylogenetic tree of select cultured Staphylococcus strains. Sequenced whole genomes were uploaded to autoMLST using de novo mode and concatenated alignment function. The resulting tree was annotated with iTOL. Type strains are colored by species. Cultured strains are in black, with # symbol indicating subject’s AD severity (none: control, #: mild AD, ##: moderate AD, ###: severe AD). (B) Heat map indicating presence of toxin genes observed in the whole genome sequences of those same selected Staphylococcus strains, as called by PROKKA. (C) In vitro enterotoxin levels in the 3 putatively unique *S. aureus* strains show strain-level differences, with SE levels increasing with severity. Bar graph depict mean and se; subject label color denotes AD severity; line represents positivity threshold.

### Effects of *Staphylococcus* supernatants on keratinocytes *in vitro*

To test the *in vitro* effects of these differences on keratinocytes, cell free conditioned media (CFCM) derived from the *Staphylococcus* isolates were incubated with keratinocytes to investigate cell viability and barrier integrity using MTT and TEER assays, respectively. Isolates from AD swabs significantly decreased cell viability (Fig 5A) and barrier function (Fig 5B). However, there was wide variability within each group.

**FIGURE 5.**
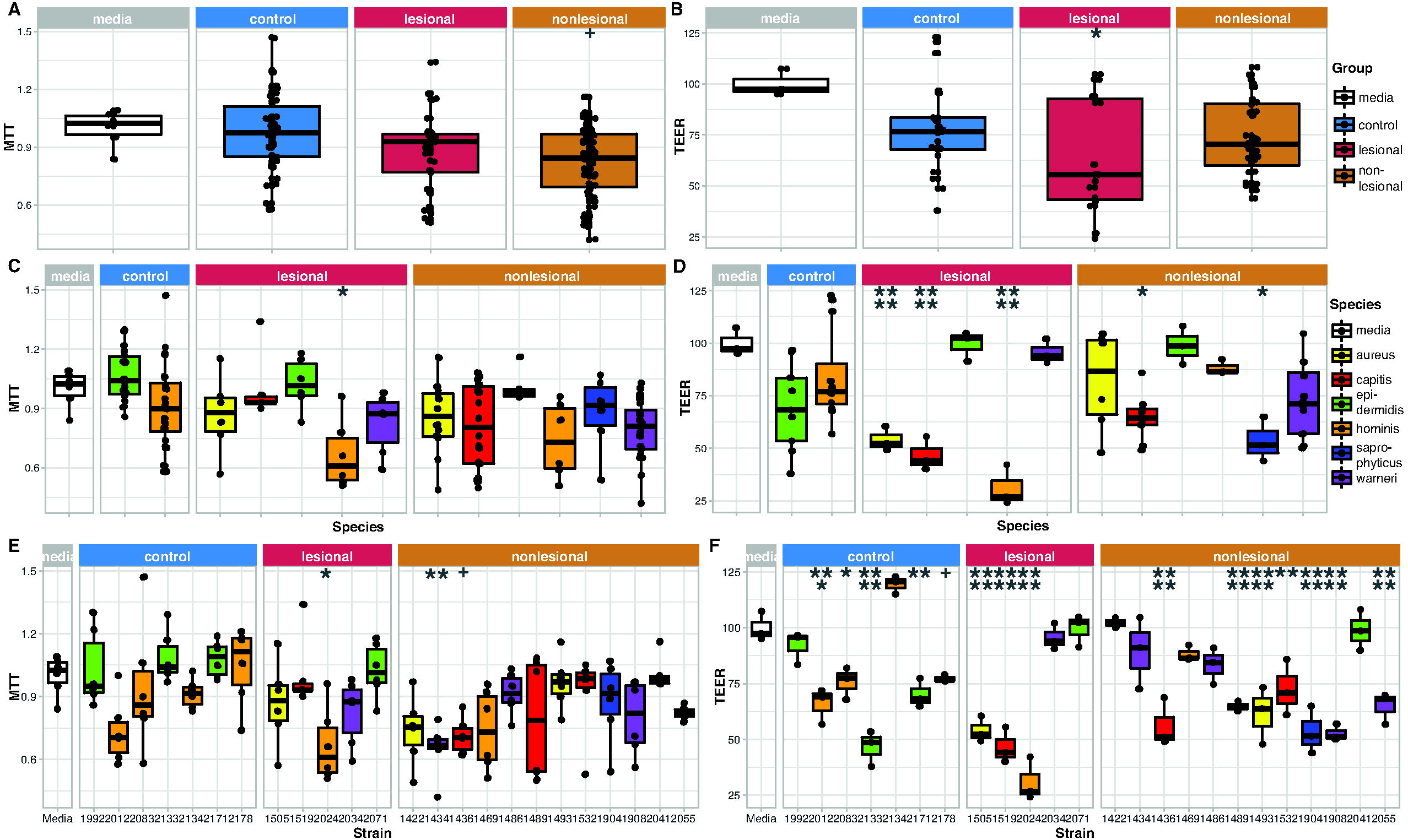
*Staphylococcus* cell-free media differentially impacts keratinocyte function. Box plots showing effects of *Staphylococcus* cell-free conditioned-media on MTT (A, C, E) and TEER (B, D, F) assays, faceted by groups and colored by species. A & B show the MTT and TEER data, respectively, at the group-level (ie all isolates in the control group are pooled, as are all in the lesional and non-lesional groups). C & D show the MTT and TEER at species-level resolution. E & F illustrate MTT and TEER with each individual strain. Bonferroni adjusted p- value symbols: + 0.15, . 0.1, * 0.05, ** 0.01, ***0.001, **** 0.0001

To investigate the variability within in each group, we first compared among species (Fig 5C, D). *S. hominis* species isolated from control swabs had no effect on cell viability, while *S. hominis* species from AD lesional swabs were detrimental. Indeed, many *Staphylococcus* species isolated from lesional swabs significantly impaired TEER (p_adj_<0.05). We next examined each isolate (Fig 5E, F). We found intriguing differences driven by the strain of Staphylococcus tested. For example, *S. hominis* LK2134, isolated from a control participant, showed a trend towards improved barrier function (p_adj_=0.12, p=0.005), whereas *S. hominis* LK2024, isolated from a lesional swab showed the most detrimental effect on barrier function (p_adj_<1E-10). *S. warneri* LK1434 significantly decreased MTT (p_adj_=0.006) but not TEER, whereas *S. warneri* LK1908 and LK2055 significantly decreased TEER (p_adj_<1E-4) but not MTT. None of the *S. epidermidis* isolates significantly impacted MTT, although 2 decreased TEER (p_adj_<0.001). All isolates of *S. capitis* significantly decreased TEER (p_adj_<0.005), but only 1 trended towards a decrease in MTT (p_adj_=0.060). Similarly, 2 strains of *S. aureus* significantly decreased TEER (p_adj_<1E-4), while the third trended towards a decrease in MTT (p_adj_=0.14).

## DISCUSSION

The skin microbiome is critical in maintaining proper functioning of the skin barrier, and previous research has focused on the role of *S. aureus* in moderate-severe AD while medications are held. In this setting, the relative abundance of *S. aureus* has been shown to increase during acute disease flares^12, 13^. In addition, *S. aureus* has also been shown to produce toxins, particularly SEB, which are associated with more severe disease^36, 37^. However, there is limited research into medication-controlled AD, which is more reflective of the clinical setting.

In our medicated cohort, we did not observe the emergence *S. aureus* blooms reported in previous studies enrolling unmedicated AD, even in lesional samples. When *S. aureus* was detected, it was primarily identified from nonlesional AD swabs. This is similar to a previous study that found mitigation of *S. aureus* overgrowth when taking medication^13^. As topical corticosteroids have been shown to decrease *S. aureus* bioburden^52^, our data together with the previous work, suggests that the interventions to manage AD are decreasing *S. aureus* overgrowth, despite disease flares.

Differences in *Staphylococcus* strains were present, including in CoNS. A strain of *S. epidermidis* (ST153) was primarily associated with healthy participants. This strain was also associated with healthy volunteers in a study of catheter-related bacteremia^53^. Conversely, metagenomes from AD lesions contained number of other *S. epidermidis* strains. Although previous studies in AD have suggested potential protective effects of *S. epidermidis*, it can behave as an opportunistic pathogen. The pathogenicity of *S. epidermidis* seems to vary greatly by strain, with certain strains being associated with health and others with disease^35, 54, 55^. Thus, these data suggest the importance of strain-level differences in both *S. aureus* and non-aureus *Staphylococcus* species in AD disease expression and severity.

In addition, we observed marked differences in toxin gene presence across the Staphylococcal isolates. All 3 *S. aureus* strains, isolated from moderate and severe participants, encoded for more toxin genes than other strains. One *S. aureus* strain (isolated from a severe patient), contained the *seb* gene and expressed this protein at high levels *in vitro*. Previous studies have suggested SEB is associated with more severe AD^36, 37^, likely due to its cytotoxic effects which disrupt barrier function^56^.

Importantly, our data evaluating keratinocyte metabolism and barrier function after exposure to secreted factors from diverse Staphylococci species demonstrates that many Staphylococcal strains are capable of disrupting keratinocyte function, even in the absence of enterotoxin production. Additionally, keratinocyte dysfunction occurred across several species, and was strain dependent. In general, *S. aureus* and *S. capitis* were associated with deleterious effects on barrier function, while *S. epidermidis* and *S. warneri* had strain-level varied impacts as measured by TEER. Finally, different strains of *S. hominis* induced opposing effects on keratinocyte integrity as measured by MTT. Ongoing research will attempt to clarify the drivers of these differential effects.

Taken together, our data underscore the importance of bacterial strain in AD disease expression and pathogenesis, and suggest that topical steroids can be effective in controlling *S. aureus* overgrowth. It is worth noting that this was a small cross-sectional study, with participants covering a range of developmental stages. The skin microbiome can change significantly from infancy through puberty^60^, and controls were age and sex matched to help address this issue. Additionally, medications used varied somewhat among participants, which may have differential impacts on the skin microbiome. Thus, future research will add additional participants to clarify these effects, and to collect additional strains of *Staphylococcus*, as well as expanding to genera beyond *Staphylococcus*.

The data presented demonstrate the critical role of strain-level differences in AD pathogenicity. Strains differed with regard to presence and amount of toxins expressed, effect on keratinocyte viability and barrier function. Additionally, deleterious effects were not exclusive to *S. aureus*. Thus, both inter- and intra-species differences impact AD pathogenesis, suggesting that strain- level differences are important considerations in AD pathogenesis and disease expression.

## Data Availability

All data produced in the present study are available upon reasonable request to the authors while waiting for public release through NCBI and SRA repositories.

## ABBREVIATIONS

AD: atopic dermatitis
AMP: antimicrobial peptides
BHI: brain heart infusion
BHIT: brain heart infusion + 0.1% Tween 80
CFCM: cell free conditioned media
CoNS: coagulase negative Staphylococci
DM: differentiation media
HK: human keratinocyte(s)
MTT: 3-(4,5-dimethylthiazol-2-yl)-2,5-diphenyltetrazolium bromide
p_adj_: adjusted p value (using Bonferroni)
SEB: staphylococcal enterotoxin B
ST: strain type
TEER: trans-epithelial electrical resistance

## ACKNOWLEDGMENTS

Our sincerest gratitude to the participants and their families who made this research possible. We would also like to thank members of both the Kalan and Singh labs for their valuable feedback during both the experimental and writing phases, especially J.Z. Alex Cheong and Elizabeth C. Townsend.

## FUNDING

NIAID K23-AI100995 (AMS)

University of Wisconsin School of Medicine and Public Health (AMS) 5T32AI007635-20 (NMLS)

NIAID U19AI142720 (LRK) NIGMS R35GM137828 (LRK)

## DATA SUMMARY

Genomic data is available on the National Center for Biotechnology Information (NCBI) database under BioProjects PRJNA830888.

## Supplemental Methods

### Patient recruitment and classification

Food allergy was diagnosed by a board certified pediatric-allergist immunologist with typical signs and symptoms of immediate Ig-E mediated reaction (ex: hives, angioedema, vomiting, *etc*.) together with evidence of allergic sensitization or oral food challenge as previously described^61^. Other allergic disease diagnosis was determined by parental report of physician diagnosis and confirmed by medical record review.

### Sample collection

Swabs were obtained from the forearm by rolling the swab with moderate pressure 10 times over the designated area, and then by turning the swab 90 degrees and rolling the swab 10 additional times. Lesional swabs were collected from sites of active lesions and nonlesionsal swabs were obtained from healthy-appearing skin from a similar body site (for example, just adjacent to the lesion, or on the contralateral body site) in a similar fashion, or from the anterior forearm when no lesion was present. Swabs were then submerged in lysis buffer and stored at -80 °C for further analysis.

### Bacterial isolation

Samples were thawed and 100 μL were pipetted onto 4 media plates—brain heart infusion (BHI) agar, BHI + 0.1% Tween 80 (BHIT) agar, blood agar, and mannitol salt agar. Glass beads were used to spread the sample evenly across plates. After the beads were removed and the plates dried, they were incubated at 28 °C for 2-3 days. At this time, morphologically unique colonies were picked from the plates and streaked for isolation onto fresh plates of the same media type. All plates were then returned to the 28 °C incubator. Every 2-3 days for 1 week, the original culture plates were checked for new colonies. An individual colony was picked from the pure cultures on the isolation plates and grown in BHIT liquid culture overnight at 37 °C. After the cultures were turbid, a 15% glycerol stock was prepared for storage at -80 °C.

Bacterial isolates from stock were grown on BHIT agar overnight at 37 °C. From these plates, one colony was inoculated into 3 mL liquid BHI + (0.1%) Tween and incubated with shaking at 37 °C overnight. The 16S rRNA gene was amplified using PCR. Briefly, 0.5 μL of the overnight culture was added to 24.5 μL of master mix (12.5 μL Econotaq PLUS green, 1 μL F primer 16S 8F (10 μM), 1 μL R Primer 16S 1492R (10 μM), 10 μL nuclease free water) and run on a thermocycler (95 °C for 10 min, [95 °C for 30 s, 54 °C for 30 s, 72 °C for 60 s] for 30 cycles, 72 °C for 5 min, hold at 4 °C). Product was confirmed by running on a 1% agarose gel and then sent for Sanger sequencing at Functional Biosciences, Inc (Madison, WI).

### Generation of staphylococcal cell free supernatants

Supplemented keratinocyte growth media was composed of 500 mL Medium 154 (Life Technologies), 500 mL Keratinocyte Serum Free Media with calcium (Life Technologies), 5 mL Human Keratinocyte Growth Serum (Life Technologies), 25 mg bovine pituitary extract (Life Technologies), 0.0025 mg human recombinant epidermal growth factor (Life Technologies), 50 mg/mL L-proline (DOT Scientific Inc), 15 mg/mL nicotinic acid (DOT Scientific Inc), and 2 mg/mL pantothenic acid (VWR)^21, 62, 63^.

### Metagenomic library preparation and analysis

Samples were processed at University of Minnesota Genomics Center (UMGC) for library preparation using the Nextera XT DNA Library Prep kit (Illumina), and metagenomic sequencing on a NovaSeq (Illumina 2 x 150 bp reads). Resulting FASTQ files were processed using quality filtering, adapter removal, human decontamination, and tandem repeat removal as described previously^40^. Taxonomic classification and abundance estimation was performed using Kraken2 (v2.0.8-beta)^41^ and Bracken (v2.5)^42^. A custom Kraken2 database was used as described previously^40^, with the addition of 165 novel skin genomes from Saheb Kashaf *et al*.^64^. Reads assigned as *Homo sapiens* or that were unclassified at the genus level were filtered out.

Decontam (v1.16.0) was then used to identify and remove contaminant reads using the “prevalence” method, based on negative controls collected at time of swab collection and processed with samples. See Supplemental Figure 2 for read counts at each step.

### MTT assay

Cell viability was assessed using the 3-(4,5-dimethylthiazol-2-yl)-2,5-diphenyltetrazolium bromide (MTT) colorimetric assay (Roche Diagnostics, Mannheim, Germany) per manufacturer instructions. For the MTT assay, 10,000 neonatal human keratinocyte (HK; obtained from American Type Culture Collection (ATCC, Manassas, Virginia) cells were plated into a 96-well plate and cultured in a 5% CO_2_ atmosphere at 37 °C. To determine the effect of the *Staphylococcus* supernatants on HK cell viability, the cells were treated with HK media alone (154 Human keratinocyte media (Thermo Fisher Scientific) supplemented with Human Keratinocyte Growth Supplement and penicillin/streptomycin (Thermo Fischer Scientific) additives) or HK media containing one of the *Staphylococcus* supernatants for 48 hours. The concentration of all supernatant treatments was standardized to 1*10^6^ CFU/mL. After the treatment period, HK were incubated in an MTT solution for 4 hours. A solubilization buffer was then added to each well and the cells were incubated in a humidified atmosphere at 37°C overnight. The absorbance was measured at 550 to 600 nm, with a reference wavelength of >650 nm, using a microplate reader (BioTek). The absorbance values were then used to determine cell viability.

**SUPPLEMENTAL TABLE 1** Table showing the medication usage and allergic co-morbidities of all subjects. AD = atopic dermatitis; FA = food allergy; F = female; M = male; N = no; Y = yes; * = irritant contact dermatitis.

**SUPPLEMENTAL TABLE 2** Table depicting the number of isolates of each species collected from each participant.

**SUPPLEMENTAL FIGURE 1** Diagram of the workflow.

**SUPPLEMENTAL FIGURE 2** Count data at each filter step.

**SUPPLEMENTAL FIGURE 3** Bar chart of the relative abundances of Eukaryotic (A) & Viral (B) species. Color and point shape correspond to AD status. *: p<0.05, **: p<0.01

**SUPPLEMENTAL FIGURE 4** Bacterial Shannon Diversity Index broken out into evenness (A) & richness (B).

