## Supplementary figures and images for "Combined metagenomic- and culture-based approaches to investigate bacterial strain-level associations with medication-controlled mild-moderate atopic dermatitis"

### Supplemental Figure 1

**Supplemental Figure 1**

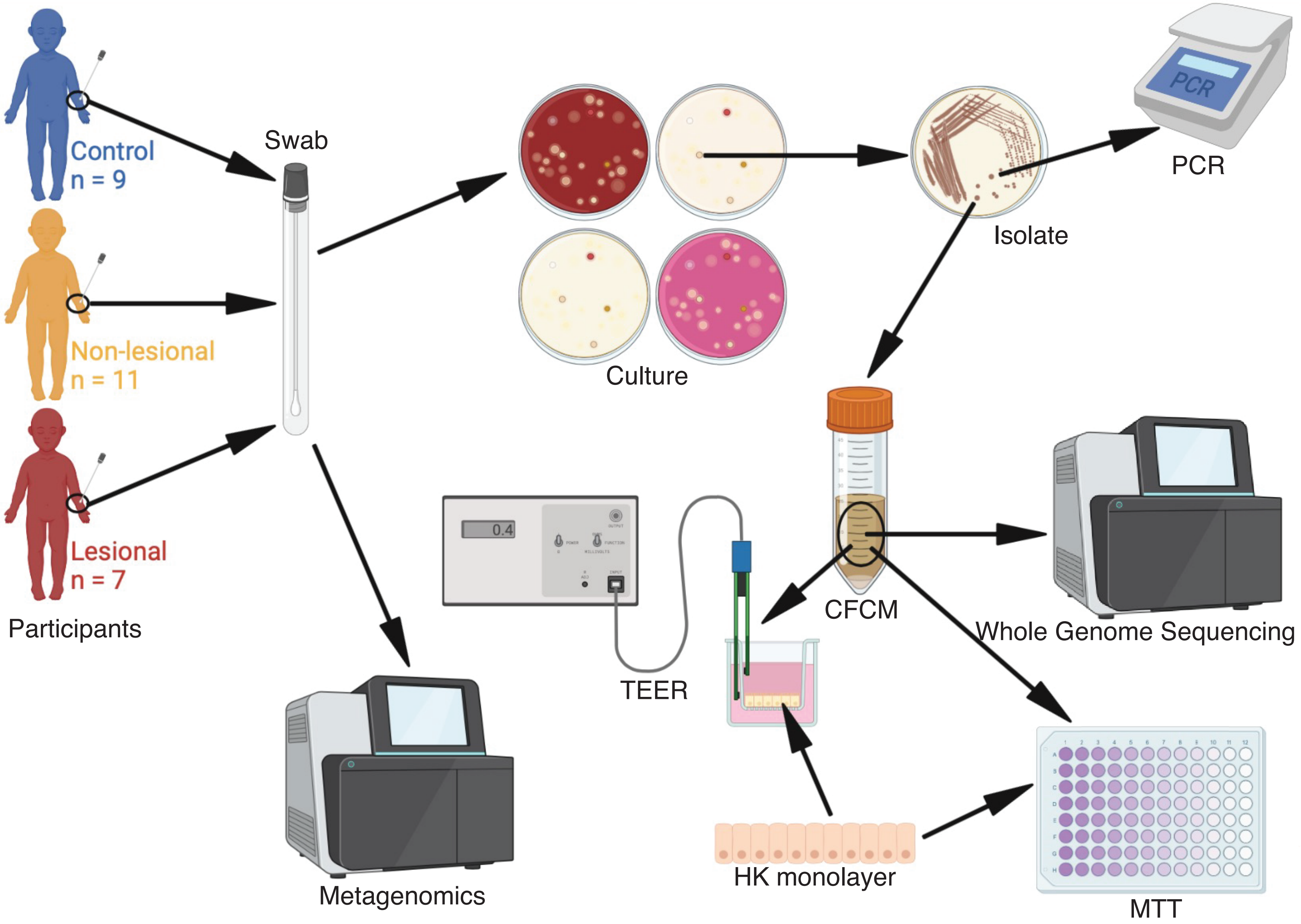

### Supplemental Figure 2

#### Supplemental Figure 2

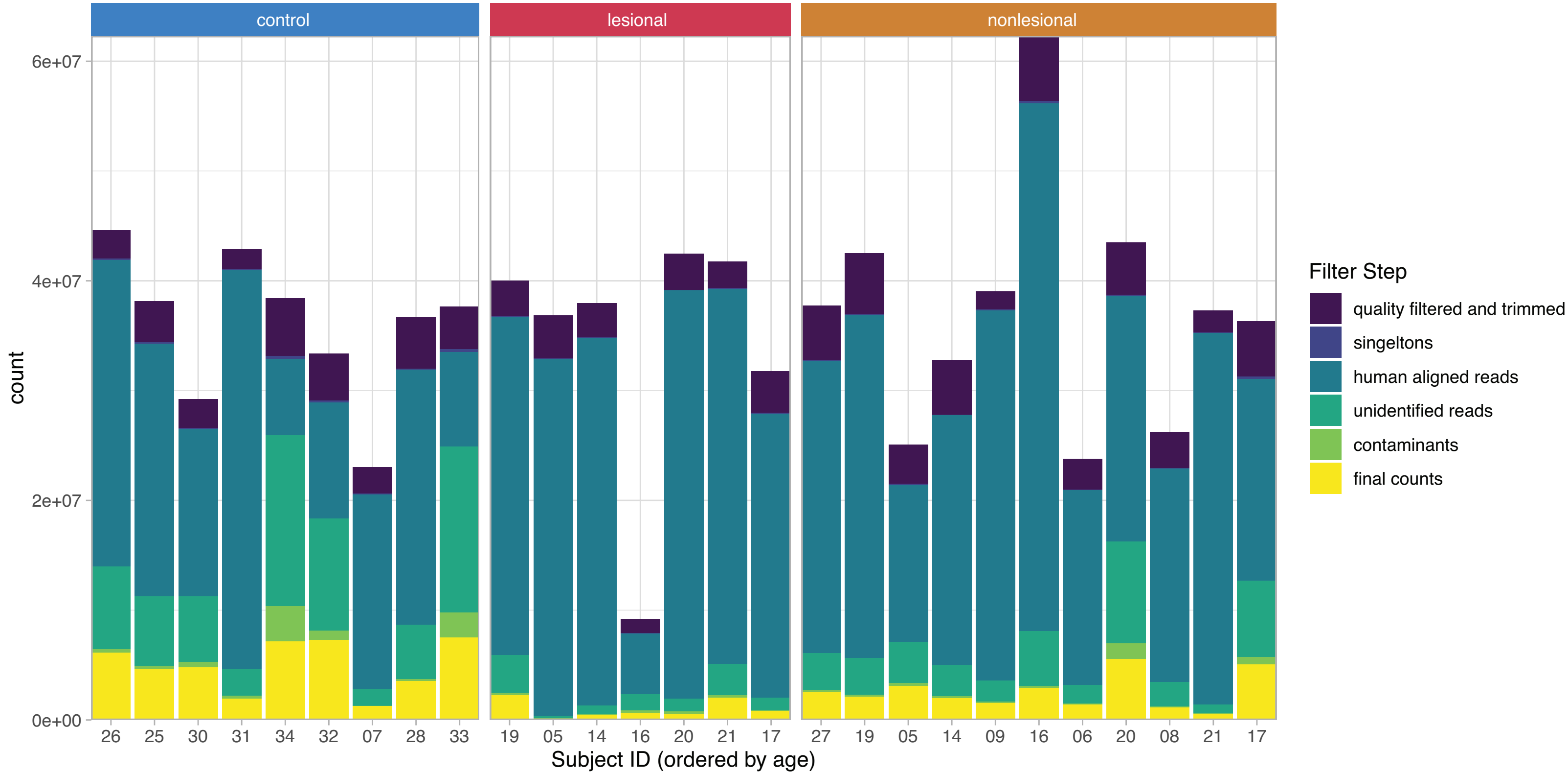

### Supplemental Figure 3

#### Supplemental Figure 3

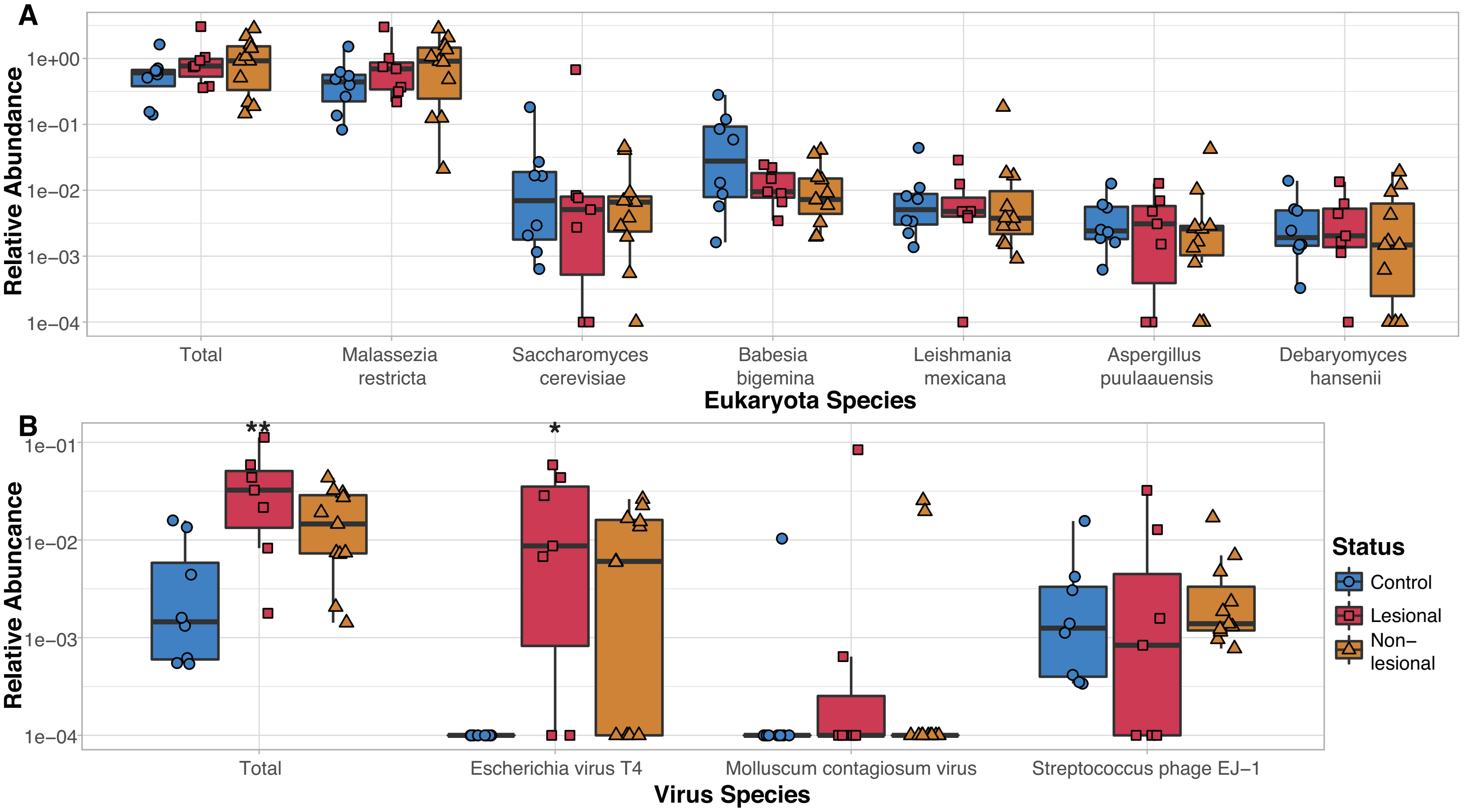

### Supplemental Figure 4

##### Supplemental Figure 4

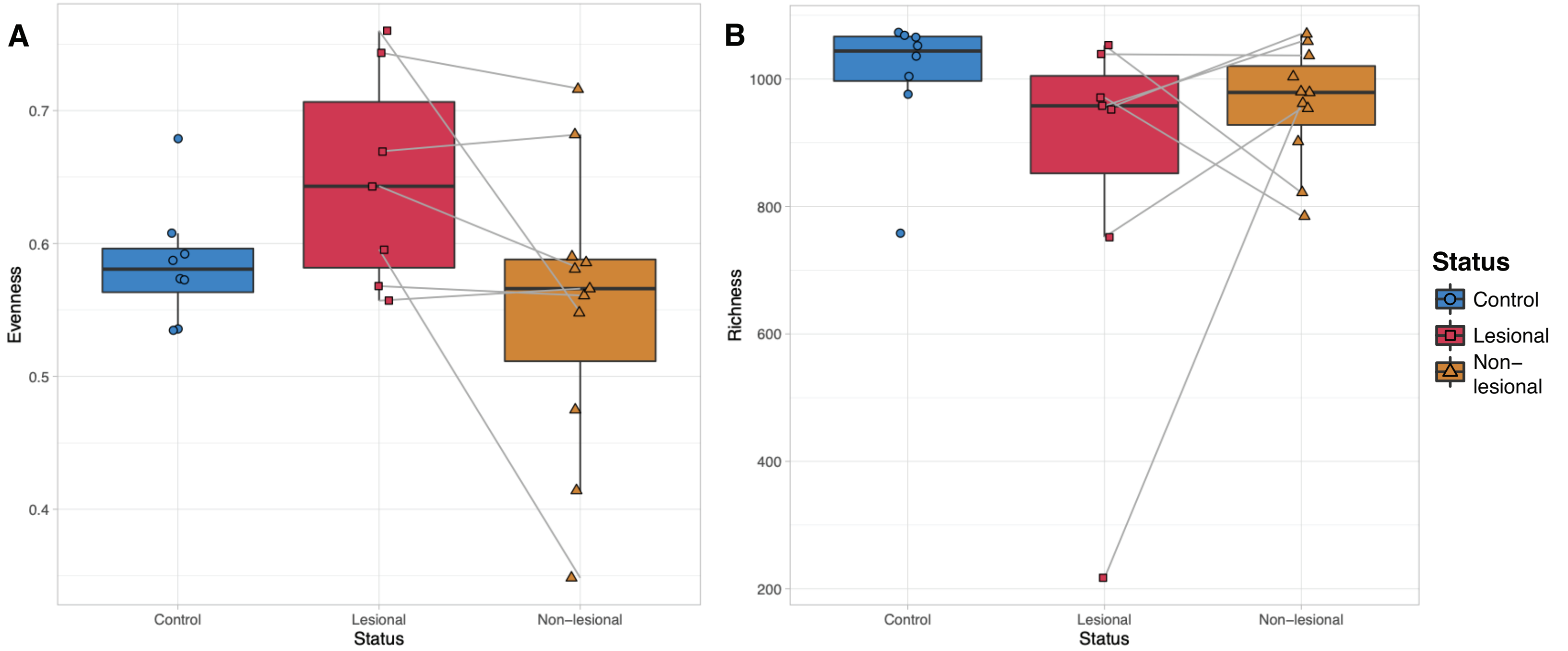
