## Supplemental Table 1 for "Combined metagenomic- and culture-based approaches to investigate bacterial strain-level associations with medication-controlled mild-moderate atopic dermatitis"

| Subject ID | Age Range | Sex | Atopic Dermatitis | Food Allergy | Family History AD | Family History FA | Asthma (med) | Rhinitis | Bleach Baths | AD Medication |
|---|---|---|---|---|---|---|---|---|---|---|
| 5 | 0-5 | F | moderate | peanut | N | N | N | N | N | protopic 0.1% ointment on face, triamcinolone 0.1% ointment (class IV) and mometasone furoate (Elocon, class II) on body |
| 6 | 0-5 | M | mild | milk | Y | N | N | N | N | triamcinolone 0.025% |
| 8 | 0-5 | M | severe | sesame | unknown | unknown | N | Y | N | hydrocortisone 2.5%, triamcinolone 0.1%, protopic |
| 9 | 0-5 | F | severe | milk, egg, peanut | Y | unknown | N | Y | Y, daily | triamcinolone 0.1% ointment (class IV), tacrolimus 0.1% ointment, hydrocortisone 2.5% |
| 14 | 0-5 | M | mild | egg (tolerant) | N | Y | N | N | N | hydrocortisone 1% |
| 16 | 0-5 | M | moderate | egg, milk, peanut, wheat, banana, beef, sunflower, green beans, tree nut | N | N | N | N | Y, 2x/wk | triamcinolone 0.1% ointment (class IV), tacrolimus 0.1% for face |
| 17 | 11-15 | F | mild | tilapia | N | N | Y (ICS) | Y | N | hydrocortisone 2.5% |
| 19 | 0-5 | F | mild | egg | Y | Y | N | N | N | triamcinolone 0.025% |
| 20 | 0-5 | M | mild | tree nuts, peanut | N | N | N | Y | N | hydrocortisone 1% ointment |
| 21 | 6-10 | M | mild | peanut, egg | N | N | Y (SABA) | N | N | hydrocortisone 2.5% |
| 25 | 0-5 | F | none | none | N | Y | N | N | N | |
| 26 | 0-5 | F | none | none | N | N | N | N | N | |
| 27 | 0-5 | M | mild | none | Y | N | N | N | N | hydrocortisone 2.5% |
| 28 | 6-10 | M | none | none | N | N | N | Y | N | |
| 30 | 0-5 | F | none | none | Y | N | N | N | N | |
| 31 | 0-5 | M | none | none | Y | N | Y (SABA) | Y | N | |
| 32 | 6-10 | M | none | none | N | N | N | Y | N | |
| 33 | 11-15 | M | none | none | Y | Y | N | N | N | |
| 34 | 0-5 | F | none* | none | N | Y | N | N | N | |
