## Supplemental Table 2 for "Combined metagenomic- and culture-based approaches to investigate bacterial strain-level associations with medication-controlled mild-moderate atopic dermatitis"

| Phylum | Genus | Control | | | | | | | | Lesional | | | | | | | Non-lesional | | | | | | | | | | | TOTAL |
|---|---|---|---|---|---|---|---|---|---|---|---|---|---|---|---|---|---|---|---|---|---|---|---|---|---|---|---|---|
| | | P26 | P25 | P30 | P31 | P34 | P32 | P28 | P33 | P19 | P05 | P14 | P16 | P20 | P21 | P17 | P27 | P19 | P05 | P14 | P09 | P16 | P06 | P20 | P08 | P21 | P17 | |
| Actinobacteria | Micrococcus | 1 | | 12 | | 9 | 5 | 38 | 24 | | 3 | 5 | | 2 | | 3 | | 2 | 2 | | 20 | 1 | 6 | 3 | 3 | 1 | 6 | 146 |
| Actinobacteria | Kocuria | | | 2 | 1 | 2 | 1 | 1 | 8 | | | 1 | 3 | | | | | | 5 | | 25 | | 17 | | | | 3 | 69 |
| Actinobacteria | Rothia | 7 | | | 6 | | | | | | | | 8 | | | | | 1 | 3 | | | | | | | | | 25 |
| Actinobacteria | Brachybacterium | | | 2 | | | | | 1 | | | | | | | 1 | | | | | | | 1 | | | | | 5 |
| Actinobacteria | Dietziaceae | | | | | | | | | | | | | | | | | | | | | | 4 | | | | 1 | 5 |
| Actinobacteria | Janibacter | | | | | | | | | | | | | | | 3 | | | | | | | | | | | 2 | 5 |
| Actinobacteria | Microbacterium | | | | 1 | | | | | | | | | 1 | | | | | | | | | | | | | | 2 |
| Actinobacteria | Brevibacterium | | | | | | | | | | | | | | | | | | 1 | | | | | | | | | 1 |
| Actinobacteria | Corynebacterium | | | | | | | | | | | | | | | | | | 1 | | | | | | | | | 1 |
| Actinobacteria | Dermacoccus | | | 1 | | | | | | | | | | | | | | | | | | | | | | | | 1 |
| Actinobacteria | Piscicoccus | | | | | | | | | | | | | 1 | | | | | | | | | | | | | | 1 |
| Firmicutes | Staphylococcus | | | 6 | 24 | 1 | | 1 | 4 | | 2 | 4 | 7 | 2 | 2 | 8 | | 9 | 13 | | 3 | 4 | 14 | 1 | 3 | | 4 | 112 |
| | S epidermidis | | | 3 | 11 | | | | 3 | | 2 | | 4 | | 2 | 2 | | 9 | 7 | | | 4 | 5 | | 1 | | 1 | 54 |
| | S hominis | | | 3 | 12 | 1 | | 1 | 1 | | | 3 | 1 | 2 | | | | | 2 | | | | 3 | | | | | 29 |
| | S capitis | | | | | | | | | | | | | | | 6 | | | 1 | | | | | | 1 | | 3 | 11 |
| | S warneri | | | | | | | | | | | 1 | | | | | | | 1 | | 1 | | 6 | 1 | | | | 10 |
| | S aureus | | | | | | | | | | | | 2 | | | | | | 2 | | | | | | 1 | | | 5 |
| | S saprophyticus | | | | | | | | | | | | | | | | | | | | 1 | | | | | | | 1 |
| | S succinus | | | | | | | | | | | | | | | | | | | | 1 | | | | | | | 1 |
| | S sp | | | | 1 | | | | | | | | | | | | | | | | | | | | | | | 1 |
| Firmicutes | Bacillus | | | | | 1 | | | 8 | | | | | | | | | | | | | | | | | | | 9 |
| Firmicutes | Streptococcus | | | | | | | | | | | | 6 | | | | | | | | | | | | | | 1 | 7 |
| Firmicutes | Enterococcus | | | | 2 | | | | | | | | | | | | | | | | | | | | | | | 2 |
| Firmicutes | Paenibacillus | | | | | | | 1 | | | | | | | | | | | 1 | | | | | | | | | 2 |
| Firmicutes | Sporosarcina | | | | | | | | 1 | | | | | | | | | | | | | | | | | | | 1 |
| Proteobacteria | Paracoccus | | | | | | | | 1 | | | | | | | 2 | | | | | | | | | | | 1 | 4 |
| Proteobacteria | Cupriavidus | | | | | | | | | | | | | | | | | | | | | 2 | | | | | | 2 |
| Proteobacteria | Enhydrobacter | | | | | | | | | | | | | | | 2 | | | | | | | | | | | | 2 |
| Proteobacteria | Sphingomonas | 2 | | | | | | | | | | | | | | | | | | | | | | | | | | 2 |
| Proteobacteria | Haematobacter | | | | | | | | | | | | | | | | | | | | | | | | | | 1 | 1 |
| | unidentified | 39 | 15 | 24 | 7 | 1 | 25 | 15 | 6 | | 1 | 7 | 9 | 1 | | 1 | 21 | 2 | 2 | | 4 | 1 | 3 | 2 | 3 | 3 | 4 | 196 |
| | TOTAL | 49 | 15 | 47 | 41 | 14 | 31 | 56 | 53 | 0 | 6 | 17 | 33 | 7 | 2 | 20 | 21 | 14 | 28 | 0 | 52 | 8 | 45 | 6 | 9 | 4 | 23 | 601 |
